# Is it safe to use a single ventilator for two or more patients?

**DOI:** 10.1101/2020.07.01.20080556

**Authors:** Sebastián Ugarte Ubiergo, Felipe Castillo Merino, Óscar Arellano-Pérez

## Abstract

Mechanical ventilation is essential in the SARS-CoV-2 pandemic context. Considering the limited availability of mechanical ventilators due to high costs increased by global demand, the use of a single ventilator for two or more patients has been encouraged. An experimental model that ventilates two test lungs with a single machine has been designed in order to measure possible asymmetries during parallel circuit ventilation under different lung compliance conditions. This paper reports a first assessment of the risks involved in ventilating two patients with a single machine. Since some volumetric differences are not monitored by the ventilator itself, the main risks involved are distension or alveolar collapse if used in actual patients that have different thoracopulmonary mechanics.

## Introduction

The SARS-CoV-2 pandemic has impacted health systems with an exponential increase in bed, professional and intensive care system demands (1). Mechanical ventilation is essential in the case of patients suffering from severe respiratory failure and require hospitalization in intensive care units (ICUs). Considering the limited availability of mechanical ventilators due to high costs, increased by global demand and based on experiences in Italy and Spain (2), sharing a single mechanical ventilator with two or more patients with has been encouraged (3).

Currently, several Scientific Societies have released statements (2) warning about potential dangers of this strategy. Their assessment is that ventilators might not be able to go beyond their initial automatic tube compensation, and volumes delivered would go to lung segments with increased compliance, PEEP could not be screened individually, pressure and volume monitoring would display the average of both patients, and each patient’s deterioration and/or recovery could occur in different time frames, among several other limitations.

Since March 16^th^, Chile has entered phase 4 of the pandemic (5). In this scenario, an experimental model has been designed to study ventilation on two test lungs with a single machine to measure possible asymmetries during parallel circuit ventilation in the case of different lung compliance.

## Method

A Puritan Bennett 840 (Covidien IIc, USA) mechanical ventilator was used, with two EasyLungTM test lungs (Imtmedical, Switzerland), each holding a compliance of 25 ml/mbar and a maximum volume of 1000 ml, 2 respirometers (Wright Haloscale, Spire), 2 pressure gauges (VBM), and 2 Disposable Ventilator Breathing Circuit Corrugated Tubes. External elastic bands were used on test lungs to increase elasticity. Ten measurements were taken on each condition; first with 2 lungs without a restrictive component (CTL) and then with one lung with a restrictive component (ITL), in volume and pressure-controlled modes. Tidal volume, maximum pressure and minute volume were measured (Table 1). Analysis was carried out using student’s t-test to determine differences between pressure-controlled and volume-controlled modes, and between the control test lung (CTL) and interventional test lung (ITL). Significance level was greater than 0.0001.

**Table 1.** compares CTL and ITL results before and after the elastic band was placed.

|  | AC/CV (Vt programmed 900 ml, Fr 20 bpm, PEEP 8 cmH2O) |  |  |  |  |  |  |  |  |  |
| --- | --- | --- | --- | --- | --- | --- | --- | --- | --- | --- |
|  | Flow (l/min) |  |  |  | Vt exhaled (ml) |  | Pressure (cmH2O) |  |  |  |
|  | Pre test | (DS) | Post test | (DS) | Pre test | Post test | Pre test | (DS) | Post test | (DS) |
| Control Test Lung (CTL) | 8540 | ±77,8 | 12902 | ±41,5 | 427 | 645,1 | 24 | ±0,5 | 47 | ±0,7 |
| Interventional Test Lung (ITL) | 9598 | ±78,9 | 4774 | ±409,7 | 478,4 | 238,7 | 24 | ±0,5 | 47 | ±1,2 |
|  | AC/CP (Pinsp. 20 cmH2O, Fr 20 bpm, PEEP 8 cmH2O) |  |  |  |  |  |  |  |  |  |
|  | Flow (l/min) |  |  |  | Vt exhaled (ml) |  | Pressure (cmH2O) |  |  |  |
|  | Pre test | (DS) | Post test | (DS) | Pre test | Post test | Pre test | (DS) | Post test | (DS) |
| Control Test Lung (CTL) | 10164 | ±5,5 | 10468 | ±494,1 | 508,2 | 523,4 | 28,4 | ±0,9 | 27,6 | ±0,5 |
| Interventional Test Lung (ITL) | 10562 | ±8,4 | 780 | ±50 | 528,1 | 39 | 28,6 | ±0,5 | 27,4 | ±0,5 |

## Results

Ventilator automatic tube compensation (ATC) was performed to evaluate pressurization and compliance of two parallel connected circuits. The test was successful.

Distensibility decrease on one of the test lungs (under identical basal conditions) ensued a smaller volume delivery than the one its counterpart got in volume-controlled mode (p <0,0001) and pressure-controlled mode (p <0,0001), while this difference was greater in pressure-controlled mode (Figure 1). System pressure increased on both circuits in volume-controlled mode. Maximum pressure difference between test lungs was not significant in either VC (p >0.9999) nor PC (p: 0.1679)

**Figure 1.**
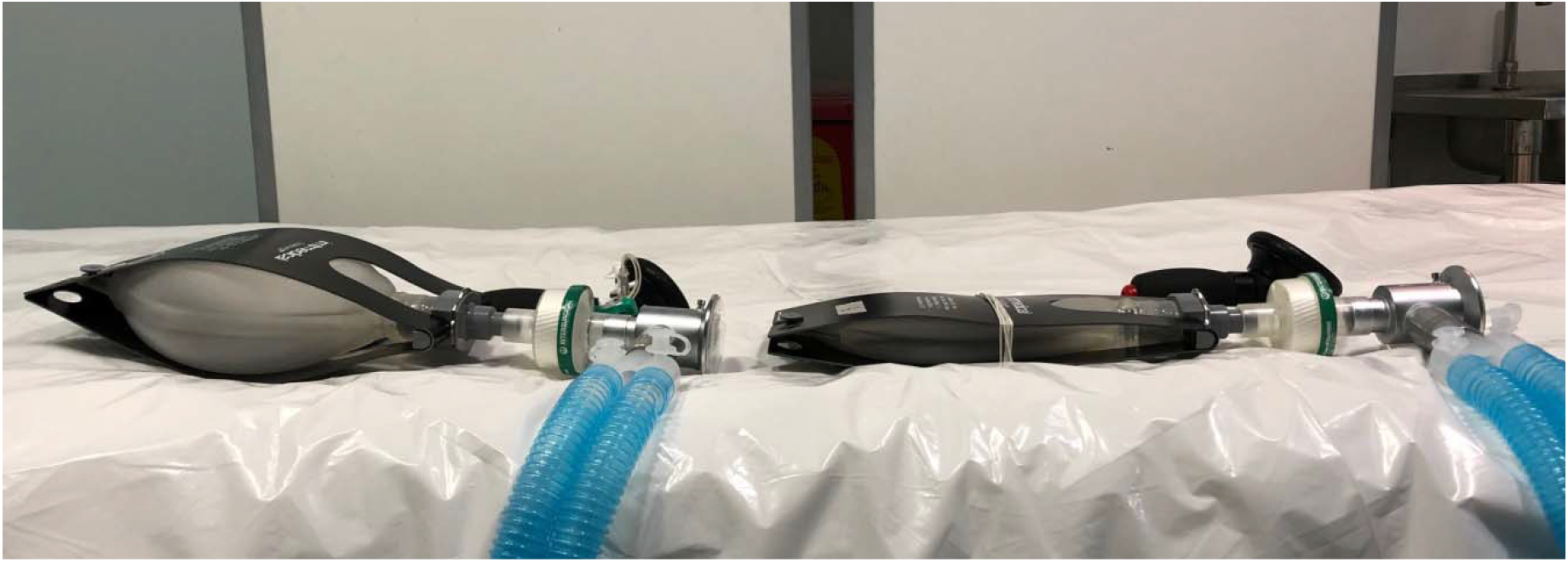

## Conclusions

The ventilator succeeded at automatic tube compensation with two parallel circuits. Tidal volumes, pressures and flows were initially similar in both modes. By adding a restrictive element to one of the test lungs, we proved that volumes given to them were different, and that maximum pressure increases in the volume-controlled mode. Volume difference showed to be greater in pressure-controlled mode. This report is a first attempt to approach the risks of ventilating two patients with a single machine. Since certain differences in volumes are not monitored by the ventilator, there are risks of distension or alveolar collapse if a single machine is used in actual patients with different thoracopulmonary mechanics (2).

## Data Availability

I declare, represent the responsibility together with the three authors, on the good practice in the writing of the manuscript to present.

